# Clinical Impact, Diagnostic Performance, and Prognostic Implications of Plasma Metagenomic Next-Generation Sequencing in Solid Organ Transplant Recipients

**DOI:** 10.64898/2026.07.02.26357172

**Authors:** Natasha Spottiswoode, Pedro S. Marra, Emily C. Lydon, Victoria T. Chu, Nathan Radakovich, Julieta Rodriguez, Hoang Van Phan, Charles R. Langelier, Monica Fung

**Affiliations:** Department of Medicine, Division of Infectious Diseases, University of California San Francisco, San Francisco, CA, USA; University of California San Francisco School of Medicine, San Francisco, CA, USA; Department of Pediatrics, Division of Infectious Disease and Global Health, University of California San Francisco, San Francisco, CA, USA; Chan Zuckerberg Biohub San Francisco, San Francisco, CA, USA

## Abstract

**Background:** Plasma metagenomic next-generation sequencing (mNGS) may detect pathogens in solid organ transplant (SOT) recipients, but optimal patient selection and result interpretation remain uncertain.

**Methods:** We studied 145 SOT recipients who received Karius plasma mNGS testing between 2017-2025. Retrospective multi-physician adjudication assessed diagnostic accuracy, clinical impact, and outcomes. We examined whether detection of atypical bacteria, invasive fungi, adenovirus/parvovirus and parasites, termed pre-specified organisms of presumed significance (POPS), predicted positive clinical impact. We applied a GPT-4o large language model (LLM) to limited electronic medical record (EMR) data to identify patients with POPS diagnoses and positive-impact testing.

**Results:** Of 145 SOT recipients, 119 (82.1%) had positive tests, and 42 (29.0%) had ≥1 POPS organism detected. Twenty-seven of 133 diagnoses (20.3%) were made by mNGS first or mNGS only. Diagnostic performance versus a gold standard of all microbiologic testing varied by organism, ranging from 100% sensitivity and specificity (*Bartonella, Nocardia*) to 60.0% and 53.3% sensitivity for *Coccidioides* and *Aspergillus,* respectively. Of 141 patients with interpretable test impact, 27 (19.1%) had positive clinical impact, associated with POPS detection (P<0.001). The LLM identified patients with POPS diagnoses (area under the receiver operating characteristic curve [AUC] 0.86), and patients with positive-impact testing (AUC 0.71).

**Conclusions:** Plasma mNGS can aid diagnosis and management of infections in SOT recipients, but negative tests do not exclude invasive fungal disease. Positive clinical impact is greatest when POPS organisms are detected, and patients at risk of POPS diagnoses may be identified by an LLM given limited EMR data.

**Summary:** Optimal use of plasma mNGS in solid organ transplant recipients remains uncertain. We find mNGS is most impactful when atypical bacteria, fungi, key viruses, or parasites are detected, and develop a method to identify patients at risk of those infections.

## Introduction

Patients who undergo solid organ transplantation (SOT) receive immunosuppressive agents that put them at risk for multiple infections^1^, including infections by fastidious or opportunistic pathogens that culture- or PCR-based techniques may miss^2^. Additionally, SOT recipients have increased infection-related morbidity and mortality compared to immunocompetent patients^3^, making prompt diagnosis urgent. Plasma metagenomic next-generation sequencing (mNGS) detects microbial DNA without culture^4,5^, potentially detecting organisms overlooked by traditional methods^6^, or missed due to culture sterilization after antibiotic administration^7,8^. However, mNGS may also produce results of uncertain significance, is relatively costly, and has limited data guiding utilization^4,9,10^. A recent single-center study of 113 SOT recipients estimated clinical benefit in 24% of patients^11^ yet left open the questions of how to target testing, clinical performance, and result interpretation.

A primary barrier to fully realizing the potential of clinical mNGS testing is that the sensitivity and negative predictive value of plasma mNGS for most pathogens and syndromes is not known, and clinicians are therefore appropriately reluctant to de-escalate empiric therapy based on negative testing. Some data exist on the diagnostic performance of plasma mNGS for *Pneumocystis jirovecii*^12^ and invasive fungal infections^13–15^, however, a large-scale, comprehensive review is needed that compares clinical plasma mNGS testing to clinically confirmed microbiological diagnoses.

Secondly, estimates of when plasma mNGS testing has a positive clinical impact vary widely, differing across patient populations, impact-assessment methods, and institutional ordering practice, and the specific test used^10,11,13,16,17^. Efforts to identify predictors of positive clinical impact for diagnostic stewardship have not succeeded in finding patient characteristics (including immunocompromised status) or clinical scenarios that reliably predict useful testing, leaving clinicians uncertain when to best deploy this test^16,18^. Specific identification of some organisms, especially atypical bacteria, invasive fungi, adenovirus/parvovirus, and parasites, have been linked with positive test impact^16,19,20^. Here, we refer to this group of high-consequence, seldom-commensal microbes as pre-specified organisms of presumed significance (POPS). However, the presence of these microbes is not known *a priori* and thus does not inform which patients should undergo testing.

Finally, the clinical significance of an mNGS test that reports multiple organisms often remains unclear: detection of multiple bacterial taxa on clinical mNGS has been linked with lower probability of positive clinical impact^16^. At the same time, research-based mNGS testing in critically ill patients with sepsis has shown that greater microbial cell free DNA burden in plasma is linked to poorer clinical outcomes, as is detection of cytomegalovirus and key pathogens such as *Staphylococcus aureus*^21^. Few studies have investigated the relationship between mNGS clinical results and patient outcomes.

Here, we study a large cohort of SOT recipients tested during routine care. We assess plasma mNGS diagnostic performance against physician-adjudicated diagnoses, show that positive impact of testing concentrates in tests that detect POPS organisms, create a tool to identify at-risk patients before testing, and examine correlations between plasma mNGS test results and patient outcomes. This study provides new insight into which patients would most benefit from mNGS and deepens our understanding of this test’s diagnostic and prognostic performance.

## Methods

### Inclusion/exclusion criteria, institutional review board approval, and data collection

We conducted a retrospective observational study of 145 patients who had undergone solid organ transplantation and had first-instance plasma mNGS testing (Karius, Inc.) at the University of California San Francisco (UCSF) Medical Center 1/2017 – 1/2025 (**Supplementary Figure 1)**. For patients with multiple plasma mNGS tests sent, only the first test sent post-transplantation was analyzed. Patient information, including demographics, transplant type and date, immunosuppression regimen, antimicrobial prophylaxis regimen, plasma mNGS results, other microbiology results, and clinical outcome were extracted from electronic medical records (EMR) in accordance with biobanking protocol #10-01116 with waiver of consent approved by the UCSF Institutional Review Board. All data were securely collected in REDCap^22^.

### Physician adjudication

Review was performed independently by 2 infectious disease physicians with full EMR access. An exact match was required for per-organism impact, primary microbiological diagnoses, and method for obtaining microbiological diagnosis (mNGS testing only, conventional microbiological testing (CMT) only, both methods but mNGS first, both methods but CMT first, or both methods simultaneously). Primary microbiological diagnoses were determined by consensus and included organisms detected by any method, including plasma mNGS, contributing to the patient’s acute illness at the time that mNGS testing was sent. All disagreements were resolved by discussion and mutual consensus, incorporating feedback from a third physician as tiebreaker if needed.

Clinical impact per organism was categorized as positive, neutral, negative, or indeterminate based on a modified set of criteria as defined by prior studies^10^ **(Supplementary Table 1).** A test with no identified organisms (negative test) could be categorized as having positive, negative, or neutral impact. As impact was assigned per organism, and not per test, a test with multiple organisms could have positive, negative, and/or neutral impacts.

### Reviewer concordance

In the 145-patient cohort, test impact could not be determined in four patients: three died before results were available and one lacked sufficient documentation to determine impact. Those four patients are excluded from clinical impact analyses **(Supplementary Figure 1)**. Of the remaining 141 patients, 115 had tests positive for any organisms, and 26 were negative. Of 115 positive tests with interpretable impact, 238 organisms were identified (10 organisms present in the 4 tests without interpretable impact). Initial reviewer assessment of per-organism impact was concordant in 187 of 238 organisms (78.6%) and divergent in 51 (21.4%); Cohen’s kappa 0.65 (95% CI 0.49-0.80, patient-clustered bootstrap). After discussion and review, 100% agreement was reached on per-organism impact, microbiological diagnoses and how diagnoses were made.

### Defining pre-specified organisms of presumed significance (POPS)

POPS organisms were pre-determined as seldom-commensal and hard-to-detect organisms that are almost always considered to result in disease when detected in plasma. POPS included bacteria that are considered atypical bacteria, invasive fungi, adenovirus/parvovirus, and parasites. A reference table of POPS organisms (**Supplementary Appendix**) was developed based on literature review, the Karius test list of Obligate/Opportunistic pathogens, and investigator consensus^16,20^.

### LLM input, scoring, and prompt engineering

To assess if artificial intelligence (AI) analysis of electronic medical record (EMR) data identifies patients at high risk of POPS and for whom mNGS testing would be useful, we used an LLM, GPT-4o at default temperature setting 0.7, implemented in Versa, a UCSF Health Insurance Portability and Accountability Act (HIPAA)-compliant model. For each patient, a single clinical note, written by the primary team on the day plasma mNGS testing was performed, was input into the GPT-4o interface. For 4 patients, a same-day note was not available; the closest possible note prior was substituted. One patient lacked available clinical notes and was excluded from analyses. Prompt engineering was carried out on four patients (2 with POPS diagnoses and 2 without), who were excluded from further analyses. In our final version of the prompt **(Supplementary Appendix)** GPT-4o was asked to provide a percentage estimate of likelihood of a POPS diagnosis. Because outputs may vary, we asked GPT-4o to estimate likelihood of POPS diagnosis for each patient three times and used the median score. The ability of GPT-4o scores to discriminate patients with POPS diagnoses and those with positive test impact was quantified using area under the receiver operating characteristic curve (AUC) with 95% confidence intervals by DeLong’s method, implemented in the pROC package in R^23^. A secondary prompt **(Supplementary Appendix)** was trialed with the same protocol.

### Statistics

Comparison of continuous variables was performed by Mann-Whitney tests. Categorical variables were compared using Fisher’s exact test given small sample sizes, with adjustment for multiple categories if more than three categories were compared. Significance was set at the adjusted P-value of less than 0.05. In the clinical outcome analysis, a logistic regression was performed. P-values less than 0.001 are shown as P < 0.001. All analysis was performed in R (The R Project for Statistical Computing, version 4.3.0). Final figure edits and assembly were performed in Adobe Illustrator (Adobe Systems, Inc.)

### Use of large language model/artificial intelligence tools

Beyond the use of GPT-4o described above, a large language model was used to assist in code and manuscript proofreading.

## Results

### Clinical and demographic characteristics of cohort

145 SOT recipients underwent first-instance plasma mNGS, including representatives of transplant recipients across major organ transplant types (**Table 1).** Tests were sent a median of 270 days after most recent organ transplant, with a range including mNGS sent on the same day as transplant (day 0) and one patient who received testing >12 years post-transplant (4,466 days). Almost all patients were receiving immunosuppression (144 of 145, 99.3%), and most tests were sent as inpatients (138 of 145, 95.2%). Inpatient tests were sent a median of 4 days after admission (range 0-89 days). The indication for testing varied, with the most common indication being pneumonia in an immunocompromised host.

**Table 1.** Clinical and demographic features of cohort by test impact. Four patients excluded who did not have interpretable test impact, including 2 lung transplant recipients, 1 heart transplant recipient, and 1 multi-organ transplant recipient. Positive test impact is defined as ≥1 organism with positive impact. P values for test indication and immunosuppression are calculated per category and adjusted for multiple comparisons, as patients may be receiving multiple immunosuppression medications at time of testing and testing may be performed for multiple reasons. Days from admission to test is calculated for inpatients only.

|  | Test with positive impact | Test without positive impact | P |
| --- | --- | --- | --- |
| **N** | 27 | 114 |  |
| *Patient Characteristics* |  |  |  |
| **Age, years** (Median, IQR) | 54.0 (41.0 - 62.0) | 57.0 (46.0 – 66.8) | 0.32 |
| **Female Sex** (No., %) | 11 (40.7%) | 44 (38.6%) | 0.83 |
| **Race** (No., %) |  |  | 0.79 |
| Asian | 6 (22.2%) | 17 (14.9%) | - |
| Black/African American | 2 (7.4%) | 7 (6.1%) | - |
| Native Hawaiian/Pacific Islander | 0 (0.0%) | 3 (2.6%) | - |
| Native American/Alaska Native | 0 (0.0%) | 2 (1.8%) | - |
| White | 9 (33.3%) | 44 (38.6%) | - |
| Other/Unknown | 10 (37.0%) | 34 (29.8%) | - |
| Multiple | 0 (0.0%) | 7 (6.1%) | - |
| **Days from transplant to test** (Median, IQR) | 479.0 (88.5 - 1270.5) | 242.5 (50.2 - 1335.2) | 0.33 |
| **Transplanted organ[s]** (No., %) |  |  | 0.47 |
| Liver | 5 (18.5%) | 25 (21.9%) | - |
| Kidney | 10 (37.0%) | 31 (27.2%) | - |
| Heart | 3 (11.1%) | 16 (14.0%) | - |
| Lung | 5 (18.5%) | 25 (21.9%) | - |
| Islet | 0 (0.0%) | 1 (0.9%) | - |
| Multi-organ | 4 (14.8%) | 16 (14.0%) | - |
| **Patient immunosuppression** |  |  |  |
| Any immunosuppression | 27 (100.0%) | 113 (99.1%) | 1.0 |
| Tacrolimus | 26 (96.3%) | 105 (92.1%) | 1.0 |
| Mycophenolate mofetil | 20 (74.1%) | 83 (72.8%) | 1.0 |
| Prednisone (>20 mg/day) | 3 (11.1%) | 27 (23.7%) | 1.0 |
| TNF inhibitor | 0 (0.0%) | 0 (0.0%) | - |
| B cell inhibitor | 0 (0.0%) | 2 (1.8%) | 1.0 |
| Thymoglobulin within 3 months | 1 (3.7%) | 10 (8.8%) | 1.0 |
| Other | 2 (7.4%) | 15 (13.2%) | 1.0 |
| *Clinical Scenario* |  |  |  |
| **Test sent as inpatient** | 27 (100.0%) | 107 (93.9%) | 0.35 |
| **Days from admission to test** (Median, IQR) | 3.0 (2.0 - 10.0) | 4.0 (2.0 - 9.0) | 0.74 |
| **Test indication** |  |  |  |
| Pneumonia in immunocompromised host | 17 (63.0%) | 56 (49.1%) | 1.0 |
| High concern for atypical infection | 10 (37.0%) | 35 (30.7%) | 1.0 |
| CNS infection | 4 (14.8%) | 17 (14.9%) | 1.0 |
| CNS infection/unable to get LP or biopsy | 0 (0.0%) | 2 (1.8%) | 1.0 |
| Deep seated infection | 6 (22.2%) | 23 (20.2%) | 1.0 |
| Fever of unclear origin | 3 (11.1%) | 27 (23.7%) | 1.0 |
| Sepsis, does not fit into above categories | 1 (3.7%) | 6 (5.3%) | 1.0 |
| Neutropenic fever; initial negative workup | 1 (3.7%) | 4 (3.5%) | 1.0 |
| Culture negative endovascular infection | 0 (0.0%) | 3 (2.6%) | 1.0 |
| Other | 1 (3.7%) | 15 (13.2%) | 1.0 |
| Unclear indication from chart review | 1 (3.7%) | 5 (4.4%) | 1.0 |
| **On antibiotics** | 23 (85.2%) | 92 (80.7%) | 0.78 |
| **Recommended by ID** (No., %) | 25 (92.6%) | 92 (80.7%) | 0.17 |
| *Test Results* |  |  |  |
| **Test positive for any organism[s]** (No., %) | 27 (100.0%) | 88 (77.2%) | **0.004** |
| **Test positive for ≥ 1 POPS organism** (No., %) | 24 (88.9%) | 16 (14.0%) | **<0.001** |

### Organisms detected by plasma mNGS testing

In 145 SOT recipients with plasma mNGS testing, 119 (82.1%) had tests that were positive for any organism. At the category level, bacteria were the most common type of microbe detected in the cohort (N = 85 patients), followed by DNA viruses (N = 46), and fungi (N = 26). The most common organisms detected were *Enterococcus* species (N = 25 patients), cytomegalovirus (CMV, N = 21) and Epstein-Barr virus (EBV, N = 14). Forty-two patients (29.0%) had ≥1 POPS organism identified by plasma mNGS, including *Aspergillus* species, agents of mucormycosis, fastidious/atypical bacteria such as *Bartonella, Nocardia,* and *Listeria* spp., and *Mycobacterium tuberculosis* (**Supplementary Figure 2)**. POPS DNA viruses identified included parvovirus and adenovirus. One patient had detection of the free-living amoeba *Balamuthia mandrillaris*.

### Diagnostic performance of plasma mNGS

At least one microbiological diagnosis was identified in 85 (58.6%) patients, with 134 diagnoses across the cohort. Excluding one St. Louis encephalitis virus case diagnosed by cerebrospinal fluid metagenomic RNA sequencing, plasma mNGS provided the first or only identification in 27 of 133 diagnoses (20.3%, **Figure 1).** Diagnostic performance varied markedly by organism (**Supplementary Tables 2, 3**). Sensitivity and specificity were high for POPS bacteria (88.2%, 97.7%, respectively) and POPS DNA viruses (100.0%, 99.3%). Diagnostic performance was lower for other bacteria (sensitivity 72.4% and specificity 62.9%, **Supplementary Table 2**), corresponding to negative and positive predictive values of 90.1% and 32.8%. Overall sensitivity and specificity for POPS fungi were 59.4% and 100.0%, respectively: 13 of 32 patients with diagnoses of POPS fungi were missed, corresponding to a negative predictive value of 89.7% across this cohort **(Supplementary Table 2)**. With respect to distinct pathogens, plasma mNGS detected all cases of *Nocardia* (6 of 6) and *Bartonella* (4 of 4) but sensitivity was lower for many fungi, detecting 6 of 8 instances of mucormycosis (75.0%), 8 of 15 *Aspergillus* (53.3%), 3 of 5 *Coccidioides* (60.0%), and 0 of 4 *Cryptococcus* (0.0%, **Supplementary Table 3**). Diagnostic testing details for all POPS cases are provided in **Supplementary Table 4**.

**Figure 1.**
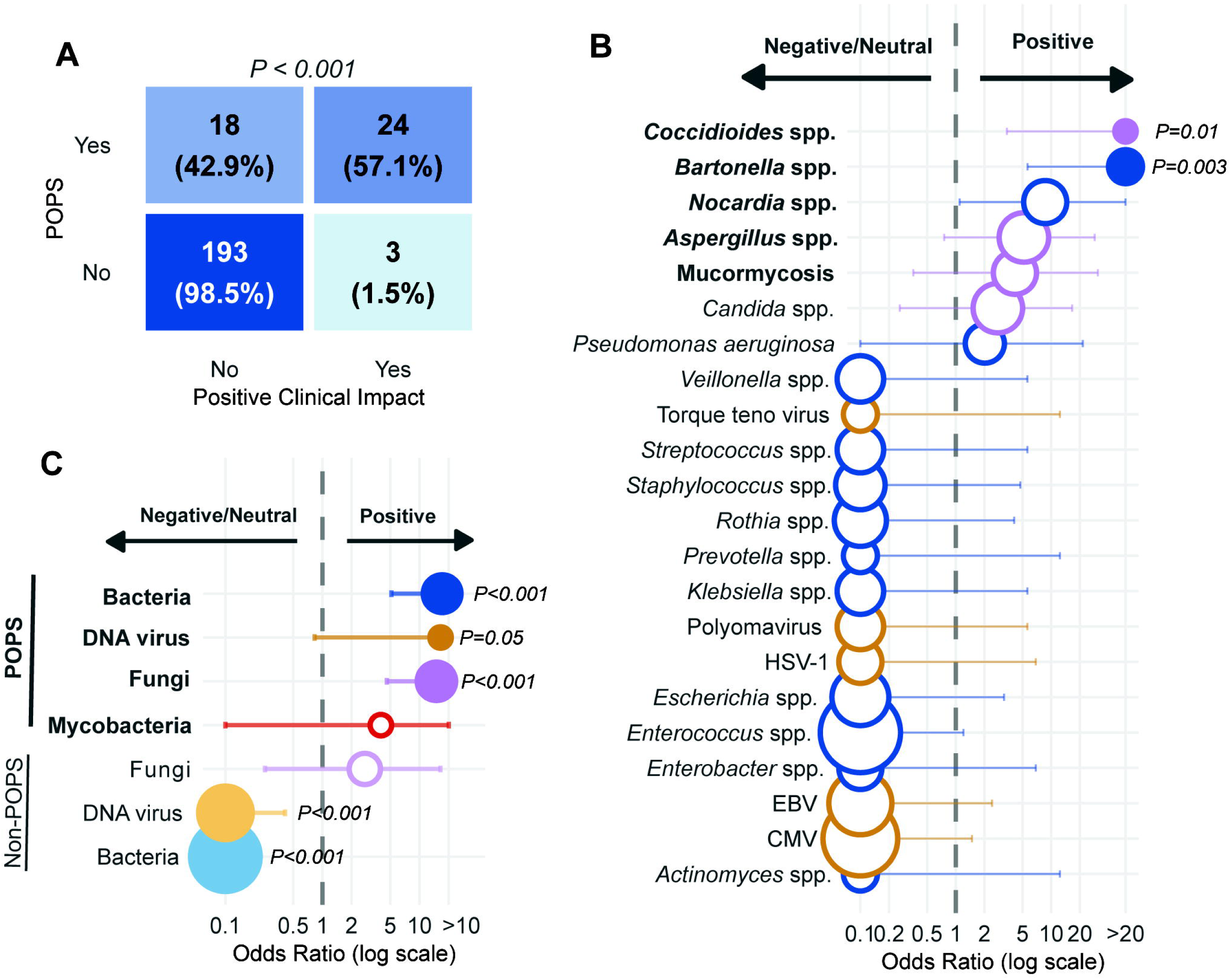
Microbiological diagnoses made by plasma mNGS versus conventional microbiology testing (CMT). Number of diagnoses in the cohort is shown on x axis; intensity of color and cross-hatching denotes how the diagnosis was made as per the legend. One patient was diagnosed with St. Louis encephalitis virus by cerebrospinal fluid RNA mNGS only; plasma metagenomic sequencing was negative and this case is excluded from the graph. Three patients contributed two diagnoses within a single genus (two patients each had two *Aspergillus* species diagnoses and one had two *Candida* species). Organisms are grouped at the genus level with the exceptions of *P. aeruginosa, S. aureus, S. pyogenes, S. pneumoniae*, and *M. tuberculosis,* as these organisms differ significantly in virulence from other bacteria in the genus; and agents of mucormycosis and non-tuberculous mycobacteria which are combined into groups. Abbreviations: mNGS, metagenomic next-generation sequencing; CMT, conventional microbiological testing; RSV, respiratory syncytial virus; HSV-1, herpes simplex virus 1; EBV, Epstein-Barr virus; CMV, cytomegalovirus; TB, tuberculosis; spp., species. Organisms categorized as POPS are shown in bold. **Figure 1 Alt Text:** Horizontal stacked bar chart listing different microbial diagnoses on the y axis, grouped by fungi, viruses, bacteria, mycobacteria, and parasites. Number of diagnoses is depicted on the x axis. Bar shading indicates how the diagnosis was made, including mNGS only, mNGS first, CMT first or simultaneous, or CMT only. POPS organisms are shown in bold text.

### Clinical impact of plasma mNGS testing

Of 141 patients for whom test impacts could be assessed, 27 tests from 27 unique patients (19.1% of total) had a positive clinical impact. Positive test impacts are itemized in **Supplementary Table 5.** Twenty-six of 27 (96.3%) positive impacts included a change in medication regimen. Nine patients had ≥1 negative impact from testing, including additional unneeded diagnostic investigations in 8 patients and additional unnecessary antimicrobial treatment in 4 patients (**Supplementary Table 6)**. A positive plasma mNGS test result was associated with a favorable clinical impact (P = 0.004, Fisher’s exact test, **Table 1**); all 26 tests negative for any organism had neutral impacts. Tests with a positive impact were most likely to have ≥1 POPS organisms detected (24 of 27, 88.9%, P < 0.001). Positive impact was not associated with patient sex, race, test indication, pre-test immunosuppression or antibiotics, or Infectious Disease involvement, though power was limited by the small number of positive-impact tests (N=27, **Table 1).** Days from transplant to test and days from admission to test did not differ between tests with and without positive clinical impact (**Table 1).** Of 7 outpatient tests, 5 were negative and 0 had a positive clinical impact.

Of the 141 patients with interpretable test impact, 238 organisms were identified. Of these, 42 were POPS organisms, identified in 40 patients. POPS status was highly associated with positive clinical impact (P<0.001, **Figure 2A**); 24 of 42 POPS organisms (57.1%) had ≥1 positive clinical impact, versus 3 of 196 non-POPS organisms (1.5%). Of the remaining 18 POPS organisms, 15 reflected confirmation of known diagnosis and 3 were considered clinically irrelevant. The three non-POPS organisms with positive impact were two detections of *Candida glabrata* and one of *Pseudomonas aeruginosa.* Of the other 193 non-POPS organisms, 128 were considered clinically irrelevant, 42 reflected known diagnoses, and 23 were associated with negative impact (**Supplementary Figure 3)**. Examining impact at the genus level, we found that plasma mNGS detection of *Coccidioides* spp. or *Bartonella* spp. was significantly associated with a positive clinical impact (both P_adj_ <0.05, **Figure 2B**). At the category level, detection of POPS bacteria, DNA viruses, or fungi was all associated with positive clinical impact (POPS bacteria and fungi both P_adj_ <0.001, DNA viruses P_adj_ 0.05). Other DNA viruses, primarily herpesviruses and torque tenoviruses, were associated with neutral/negative clinical impact, as were other bacteria (both P_adj_ <0.001, **Figure 2C**). The single parasite detected by plasma mNGS in this dataset, *Balamuthia mandrillaris*, had a positive clinical impact.

**Figure 2.**
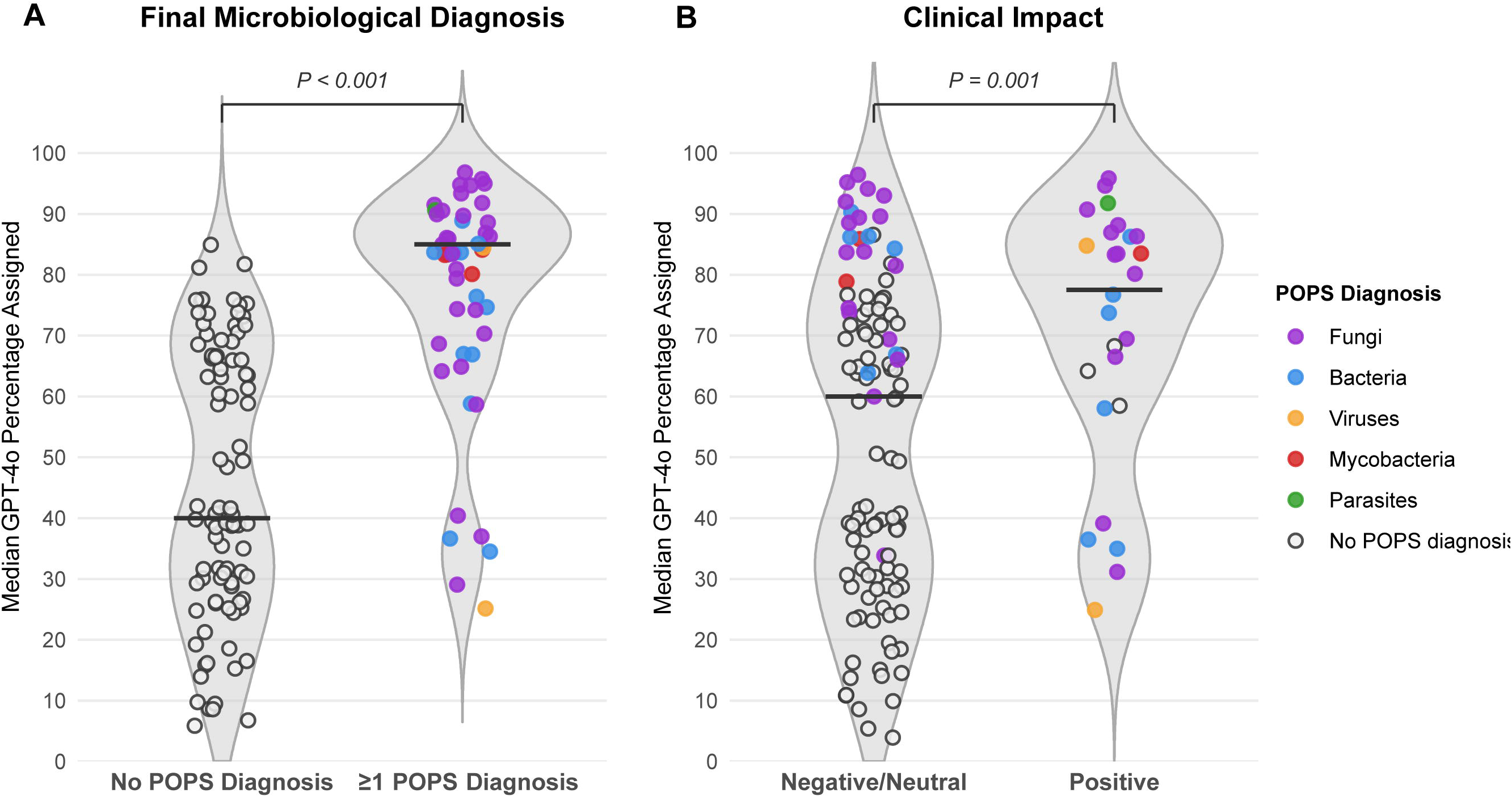
Clinical impact of plasma mNGS testing by organism. **(A)** Relationship between organism status as POPS and positive clinical impact. **(B)** Organism detected as compared to per-organism impact. X axis shows odds ratio of positive impact versus neutral/negative. Organisms are grouped at the genus level with the exceptions of *P. aeruginosa, S. aureus, S. pyogenes, S. pneumoniae*, and *M. tuberculosis,* as these organisms differ significantly in virulence from other bacteria in the genus; and agents of mucormycosis and non-tuberculous mycobacteria which are combined into groups. Only organisms detected in 3 or more tests are included, and POPS organisms are bolded. **(C)** Organism category detected compared to clinical impact. Parasites are excluded as only 1 was detected in this cohort. All odds ratio testing are Fisher’s exact tests, panels B and C include Benjamini-Hochberg correction for multiple comparisons. In **(B-C),** filled circles are significant at threshold of P<0.05 after correction for multiple comparisons, and size of circle is proportional to the number of times organism were detected in this cohort. Abbreviations: POPS, pre-specified organisms of presumed significance; spp; species; HSV-1, herpes simplex virus 1; EBV, Epstein-Barr virus; CMV, cytomegalovirus. Colors: purple, fungi; blue, bacteria; gold, virus; red, mycobacteria. N = 141 patients with interpretable clinical impact. **Figure 2 Alt Text:** Three-panel figure on clinical impact by organism. Panel A shows a two by two heatmap of POPS status (rows) versus positive clinical impact (columns) with cell counts and percentages. Panels B and C are forest plots of odds ratios of positive impact versus negative/neutral impact on a log scale x-axis, with a dashed vertical line showing an odds ratio of one. Panel B shows odds ratio by organisms grouped by genus, while panel C shows odds ratio by organism category. The size of each point in panels B and C is proportional to how often the organisms were detected; filled points mark statistical significance.

### LLM analysis of EMR data to identify patients likely to benefit from plasma mNGS testing

We created a GPT-4o prompt in a HIPAA-compliant tool that estimates a probability of POPS diagnosis based on a single note from the EMR. Excluding four patients for prompt engineering and one lacking available EMR data (**Supplementary Figure 1),** our prompt assigned higher probability of a POPS diagnosis in patients with final diagnoses of POPS organisms (**Figure 3A**, AUC 0.86, 95% CI 0.80-0.93, median GPT-4o score 85.0% versus 40.0%, P<0.001). Further excluding the 4 patients with uninterpretable test impact, we found the same prompt assigned a higher probability to patients who had tests with positive impact versus those who had tests with negative or neutral impact (**Figure 3B**, AUC 0.71, 95% CI 0.60-0.81, median GPT-4o score 77.5% versus 60.0%, P = 0.001). Had plasma mNGS testing been sent only in patients with a threshold GPT-4o median score of 30% or greater, 25 of 26 of the patients with a positive test impact would have been tested (96.2%), and 21 of 110 (19.1%) tests with negative/neutral impact would have been avoided. To ascertain if the explicit consideration of POPS diagnoses is needed for LLM prediction of test impact, we also trialed a prompt designed to predict positive impact directly. This approach failed to distinguish patients with positive-impact testing from those with negative or neutral testing (**Supplementary Figure 4**).

**Figure 3.**
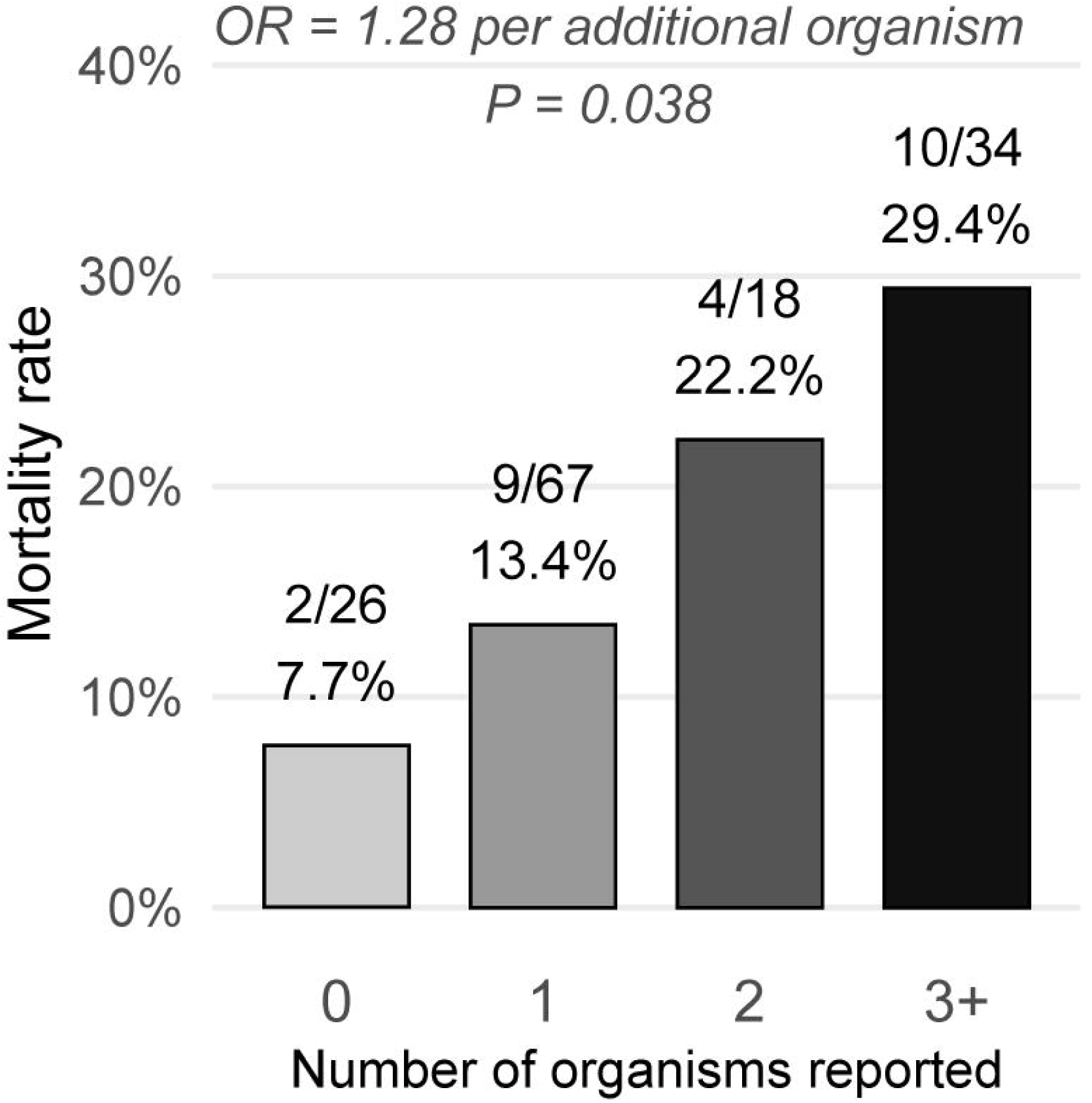
LLM analysis of EMR data identifies patients with POPS diagnoses and who had positive-impact plasma mNGS testing. **(A)** Violin plot showing GPT-4o scores in patients with final microbiological diagnoses that included POPS organisms or did not include POPS organisms. **(B)** Violin plot showing GPT-4o scores in patients with positive impact mNGS testing versus negative/neutral impact. P values are Mann-Whitney tests. Every dot is a unique patient and test; color of dots in the POPS-positive organisms indicates the type of organism detected. For 6 patients with multiple classes of POPS, the following hierarchy was used to determine color: fungi> mycobacteria> bacteria> virus. **Figure 3 Alt Text:** Two violin plots are shown. Median GPT-4o score is shown on the y axis (0- 100%). Panel A compares patients grouped by final microbiological diagnosis (no POPS diagnosis versus one or more POPS diagnoses). Panel B compares patients grouped by clinical impact (positive impact versus negative/neutral). Each dot is one patient and is colored by the POPS diagnosis, with open circles for patients without a POPS diagnosis.

### Prognostic value of plasma mNGS

Finally, we assessed the relationship between plasma mNGS results and clinical outcomes. In this dataset, 25 of 145 patients (17.2%) had a clinical outcome of mortality, as defined by death in hospital or within 30 days of discharge, or within 30 days of plasma mNGS for the 7 patients tested as outpatients. A higher number of organisms detected by plasma mNGS was associated with mortality, agnostic to POPS status (unadjusted OR = 1.28 per additional organism, 95% CI 1.01-1.61, P=0.038, **Figure 4).** No single organism, category of organisms, or POPS presence/absence was significantly associated with mortality (**Supplementary Figure 5).**

**Figure 4.**
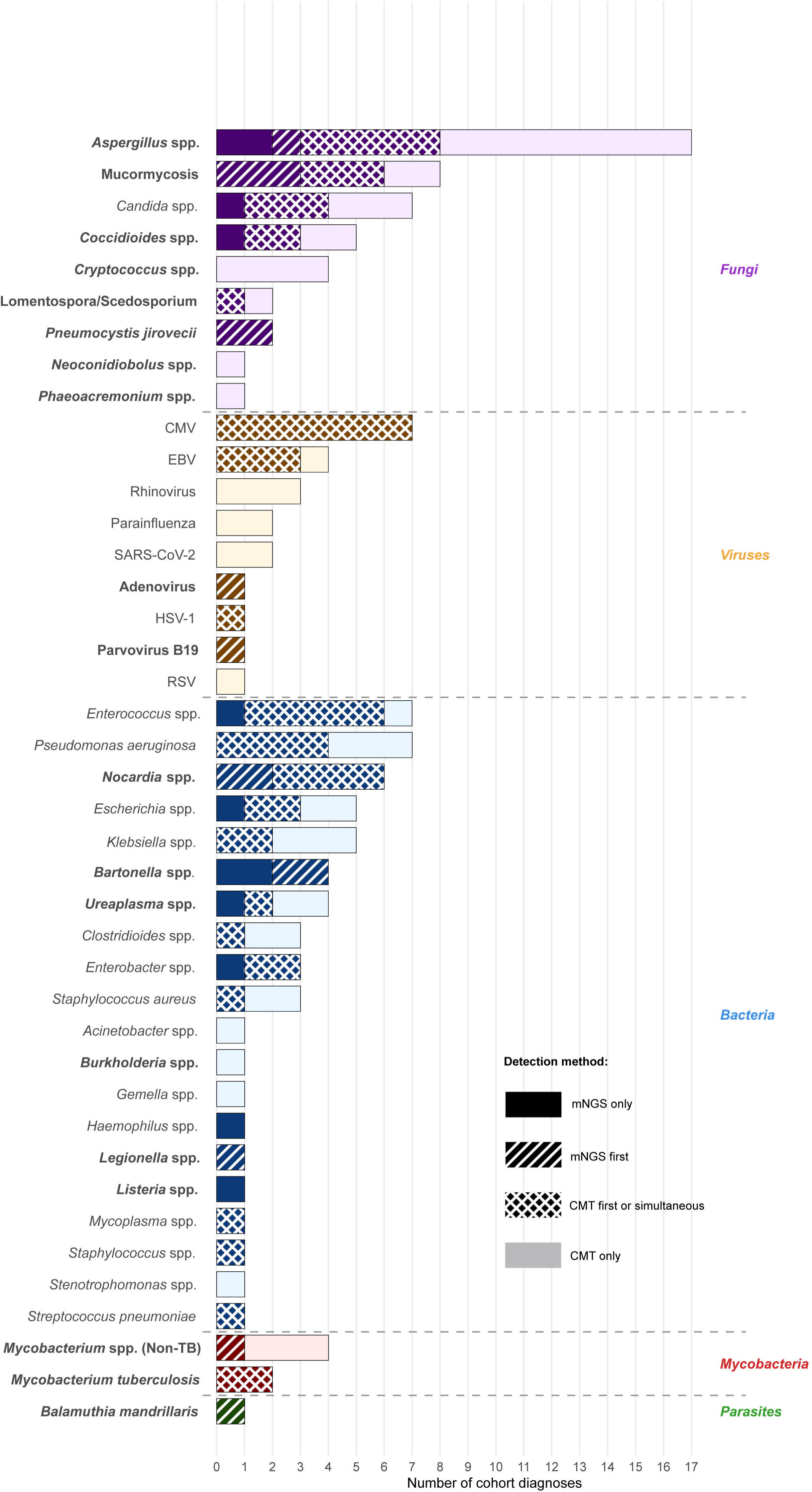
Relationship between number of organisms detected by plasma mNGS and patient mortality. Analysis based on logistic regression. OR = odds ratio per organism. **Figure 4 Alt Text**: Bar chart showing number of organisms detected by plasma mNGS (0, 1, 2 or 3 or more organisms) on the x axis. Mortality is shown on the y axis.

## Discussion

We performed a retrospective analysis on 145 SOT recipients who received clinical plasma mNGS testing in a major U.S. academic medical center from 2017 to 2025. This is the largest plasma mNGS analysis to date in this important population and the first to propose a decision tool to identify patients most likely to benefit from testing. We characterize diagnostic performance by organism, show that clinical impact concentrates in a defined set of pre-specified high-virulence and seldom-commensal pathogens, build a HIPAA-compliant LLM tool to identify patients at risk of these infections before testing, and show that organism burden carries prognostic as well as diagnostic information.

Of the microbiological diagnoses in this cohort, one-fifth were made by plasma mNGS only or mNGS first. The sensitivity and specificity of mNGS testing varied significantly, with excellent test characteristics for atypical bacteria such as *Nocardia* or *Bartonella,* but much more mixed performance in other bacteria, with a positive predictive value of 32.8%. Critically, our work confirms that plasma mNGS is not sensitive enough to serve as a rule-out test for invasive fungi. We found a sensitivity of only 53.3% for *Aspergillus*, similar to what has been reported in other cohorts^13–15^, and lower sensitivity for other key fungi, corresponding to an overall negative predictive value of 89.7%.

A recent study found that positive tests that identify atypical bacteria, fungi, parvovirus/adenovirus, and parasites (here termed POPS) are associated with a positive impact^16^, a finding that was reflected in our study. Indeed, the two organisms most strongly associated with positive clinical impact, *Coccidioides* and *Bartonella,* are almost always considered causative when identified, but are typically diagnosed by antibody-based tests^24^, which may exhibit diminished sensitivity in immunocompromised hosts^25^. As POPS organisms are almost always interpreted as causing disease when detected, their association with positive impact could be considered definitional; but two observations argue against circularity. Firstly, almost half of POPS detections produced no impact because they confirmed an already-established diagnosis. Secondly, only 3 of 196 detected non-POPS organisms produced positive impacts, indicating that utility signal of mNGS is truly concentrated in POPS detections rather than a definitional artifact.

As the most impactful mNGS test results identified POPS organisms, we developed a tool to identify patients at risk of these organisms to prioritize for testing. The GPT-4o prompt we created identifies patients with ultimate diagnoses of POPS organisms and who were more likely to have positive impact mNGS testing. At a permissive scoring cutoff appropriate for this high-risk cohort, approximately one-fifth of the lowest-yield tests could have been avoided, while still capturing all but one positive impact test. The model read the clinical notes written the day testing was performed, and so is best understood as a tool to extract and calibrate clinical information and diagnostic concern latent in the record. Although this analysis is a proof-of-concept, the possibility of complementing expensive diagnostics with a simple, low-cost LLM decision-assist tool is attractive. Our prompt is publicly available and may be freely used by any clinician with access to a HIPAA-compliant chatbot interface.

Finally, we noted that the number of organisms detected by plasma mNGS was associated with mortality (unadjusted OR 1.28, 95% CI 1.01-1.61). We hypothesize that organism burden may reflect severity of illness, related to the immune dysfunction of critical illness and/or the loss of gut barrier integrity, consistent with prior studies that link mortality to detection of cytomegalovirus DNA^26,27^, as well as to plasma bacterial DNA burden^21^.

Strengths of this study include that this is the largest plasma mNGS study to date of SOT recipients, with comprehensive multi-physician review of per-organism impact and final microbiological diagnoses. Moreover, this study is the first to leverage AI-enabled EMR analysis towards mNGS diagnostic stewardship. Weaknesses of this study include its retrospective, single-center nature; additionally, the small number of certain pathogens limit conclusions regarding sensitivity and specificity. Future prospective studies should incorporate LLM-assisted patient selection for mNGS use to fully realize the clinical potential of plasma mNGS in SOT recipients and other at-risk populations.

In conclusion, we find that in SOT recipients, plasma mNGS is most useful when the patient and clinical scenario raise high concern for atypical bacteria, fungi, adenovirus/parvovirus, or parasites, that a negative result should not be used to exclude invasive fungal disease, and that positive test impacts are concentrated enough that pre-test triage, including AI-enabled triage from limited EMR data, is feasible.

## Data Availability

All patient-level demographic data and test result data, in deidentified and HIPAA-compliant form, are available in a GitHub repository with this paper at https://github.com/infectiousdisease-langelier-lab/SolidOrganTransplant_mNGSAnalyses.git

## Acknowledgements

Conceptualization: N.S., M.F., E.C.L., V.T.C., and C.R.L.

Methodology: N.S.

Validation: H.V.P

Analysis: N.S.

Investigation: N.S., E.C.L., V.T.C., N.R., J.R., M.F.

Data curation: N.S.

Writing – Original Draft: N.S. and P.S.M.

Writing – Review and Editing: N.S., P.S.M, E.C.L., V.T.C., N.R., J.R., C.R.L., M.F.

Supervision: M.F. and C.R.L

The authors wish to acknowledge the intellectual contributions by Dr. Charles Y. Chiu. Source data are provided with this paper. All patient-level demographic data and test result data, in deidentified and HIPAA-compliant form, are available in a GitHub repository with this paper at https://github.com/infectiousdisease-langelier-lab/SolidOrganTransplant_mNGSAnalyses.git. Large language model/artificial intelligence tools were used in the GPT-4o prediction analysis as specified in Methods, using the HIPAA-compliant GPT-4o version implemented in UCSF Versa. Further, a large language model was used to assist in code and manuscript proofreading.

