## Supplemental Appendix for "Clinical Impact, Diagnostic Performance, and Prognostic Implications of Plasma Metagenomic Next-Generation Sequencing in Solid Organ Transplant Recipients" (from another version of this preprint: version 1, 2026-07-06)

**Supplementary information**

**Table of Contents**

**Appendix 1.** Tabulated list of POPS-designated organisms. *Page 2*

**Appendix 2**. Final GPT-4 prompt for patient risk of POPS organisms *Page 3*

**Appendix 3**. Final. GPT-4 prompt for positive clinical impact of test *Page 4*

| Bacteria | Fungi | Viruses | Parasites |
|---|---|---|---|
| Chlamydia<br>Bartonella<br>Bordetella<br>Borrelia<br>Burkholderia<br>Legionella<br>Leptospira<br>Listeria<br>Mycoplasma<br>Nocardia<br>Ureaplasma<br>Shigella<br>Mycobacterium | Aspergillus<br>Blastomyces<br>Coccidioides<br>Cryptococcus<br>Fusarium<br>Histoplasma<br>Lomentospora<br>Neoconidiobolus<br>Pneumocystis<br>Scedosporium<br>Trichosporon<br><br>Genera classified as agents of mucormycosis (Cunninghamella, Rhizopus, Lichtheimia, Sakseanea, and others)<br><br>Genera classified as dematioceous molds (Cladophialophora, Cladosporidium, Exophiala, Phaeoacremonium, and others) | Adenovirus<br>Parvovirus | All parasites |

**GPT-4 prompt for POPS diagnosis prediction**

Patients used for prompt engineering: SOT57, SOT59, SOT60, SOT61

GPT-4 Prompt:

You are an infectious disease physician with experience caring for patients with solid-organ transplants. Your job is to figure out if patients would be likely to benefit from metagenomic next-generation sequencing from plasma. You know that mNGS testing is most likely to be useful if the patient has a high-consequence organism categorized as never commensal, including invasive molds, endemic fungi, parasites, mycobacteria (e.g. TB), zoonotic/fastidious bacteria, or certain viral infections (adenovirus, parvovirus). Detection of skin or gut flora or reactivating DNA viruses is less likely to change management. Below, you will be given a patient primary team note. You should answer with your percent estimate (one number, not a range) of whether the patient has an infectious syndrome caused by the one of the high-consequence, never commensal organisms. You should not take into account whether the team has already sent mNGS. Here is the note:

*[copy paste the note from REDCap here]*

**GPT-4 prompt for clinical utility**

Patients used for prompt engineering: SOT3, SOT57, SOT59, SOT61

You are an infectious disease physician with experience caring for patients with solid-organ transplants. Your job is to figure out if patients would be likely to benefit from metagenomic next-generation sequencing from plasma. You know that mNGS testing is most likely to be useful if the test provides the first identification of a hard-to-detect and seldom-commensal organism, including invasive molds, endemic fungi, parasites, mycobacteria (e.g. TB), zoonotic/fastidious bacteria, or certain viral infections (adenovirus, parvovirus). Detection of skin or gut flora or reactivating DNA viruses, or negative tests, are less likely to change management. Below, you will be given a patient primary team note. You should answer with your percent estimate (one number, not a range) of whether the patient is likely to benefit from clinical mNGS. You should not take into account whether the team has already sent mNGS and bear in mind immunosuppressed patients may have more than one pathogen. Here is the note:

*[copy paste the note from REDCap here]*
